# Needlestick Injuries in Healthcare, Research and Veterinary Environments in a Sample Population in British Columbia and their Economic, Psychological and Workplace Impacts

**DOI:** 10.1101/2023.04.27.23288880

**Authors:** Jamie Magrill, Sharon Yuen Sa Low, Ina Na

## Abstract

**Introduction:** Needle-stick injuries (NSIs) are defined as the sharp point of a needle puncturing human skin. This article examines the risk and illustrates the burden of NSIs for workers in the healthcare, veterinary and research industries, and includes a sample survey population of workers in workplaces using needles.

**Methods:** For the review component of this article, PubMed and Google Scholar were queried within the date range of 1998-2022, retrieving 1,437 results. A publicly available sample population dataset was and analyzed from British Columbia (n=30) on workplace needlestick injuries. The OSHA, WHO, and NIEHS guidelines were reviewed, and the WorkSafe BC injury database was searched using FIPPA requests.

**Discussion:** Recapping remains a common practice despite decades of guidelines recommending against recapping. NSI research is underpowered and underrepresented in non-healthcare settings. NSIs lead to heightened anxiety, depression, and PTSD in workers and exposure to pathogens, toxic chemicals and permanent tissue damage. NSI annual reporting is likely an underestimate due to chronic underreporting, and the financial impact including work-loss and healthcare costs continues to rise. Current NSI prevention devices have limited uptake and thus, more affordable, versatile and efficient NSI-prevention devices are needed.

**Relevance:** Due to COVID-19, healthcare workers are at a higher risk of receiving NSIs. Emphasis on safe needle handling practices is necessary to maintain workers physical and psychological safety, to protect workers using COVID-19 PPE on long shifts, and to deliver the high volume of vaccinations required to inoculate the global population.

**Conclusion:** NSIs are detrimental to healthcare workers wellbeing, chronically underreported, and poorly surveyed. Areas of future research include determining more effective solutions to reduce NSIs, assessing the validity of NSI reporting systems, and integrating solutions with COVID-19 prevention and vaccination protocols.

## Introduction

Needlestick injuries (NSIs) occur frequently in fast-paced environments and are considered a significant occupational hazard for millions of workers including nurses, clinicians, laboratory technicians, surgeons, veterinarians, veterinary technicians, and research assistants who use needles on a daily basis (1-2). An NSI is defined as a tip of a syringe needle or other sharp object unintentionally puncturing human skin (3). Common types of needles resulting in NSIs range from hypodermic needles (32%), suture needles (19%), winged steel needles (12%), IV catheter stylets (6%), to phlebotomy needles (3%) (Fig. 1) (4). While performing an injection, users are at-risk of receiving an NSI on up to four occasions: while uncapping to draw down fluid, recapping to maintain sterility until it is time to perform the injection, uncapping to perform an injection, and recapping prior to disposal (Fig. 2) (5,6). For healthcare workers who receive an NSI, this can lead to the transmission of blood-borne pathogens such as hepatitis B and C, and HIV along with significant psychological side effects (7). Similarly, veterinary workers who received an NSI have reported experiencing severe allergic reactions, localised necrosis, abscess formation, joint infection, local nerve damage, bronchial and laryngeal spasm, blastomycosis, skin slough, and psychedelic experiences (8). The COVID-19 pandemic has led to an increase in mandatory vaccinations, increased strain on the global healthcare system, and a rise in healthcare worker burnout (9). In the veterinary field, the prevalence of global lockdowns has led to an increasing global interest in pet adoption, and thus more mandatory pet vaccinations (10). This has put additional pressure on healthcare and veterinary workers who are more likely to receive an NSI while performing repetitive medical procedures, such as vaccination and repeat injections due to an increased workload and a high-stress environment (11).

**Figure 1.**
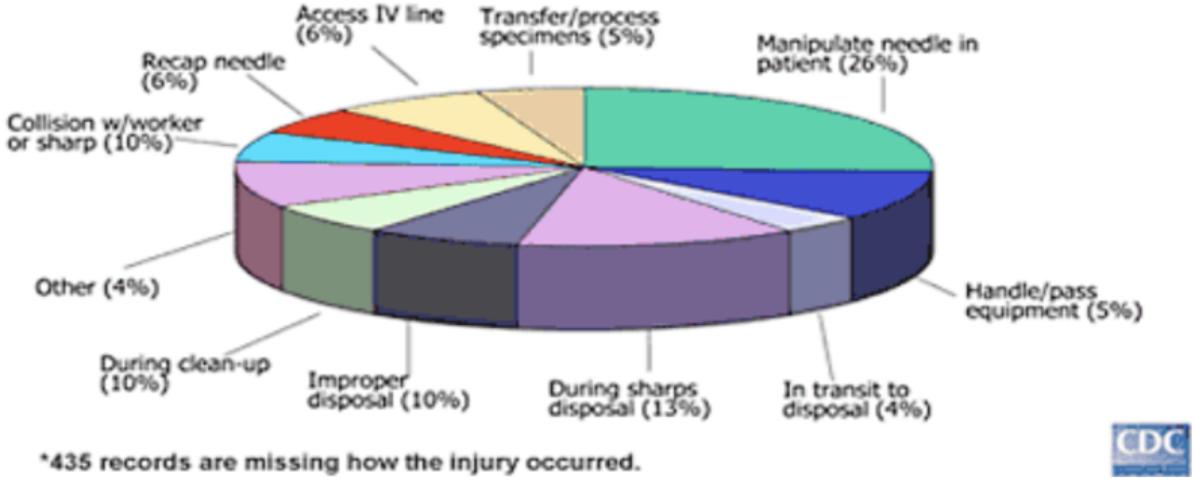
A breakdown of NSI occurrences involving hollow-bore needles (4, p.8). The main factors that influence the frequency of NSI are equipment design, nature of the procedure, work environment, staff experience, recapping and disposal of needles. Reproduced with permission.

**Figure 2.**
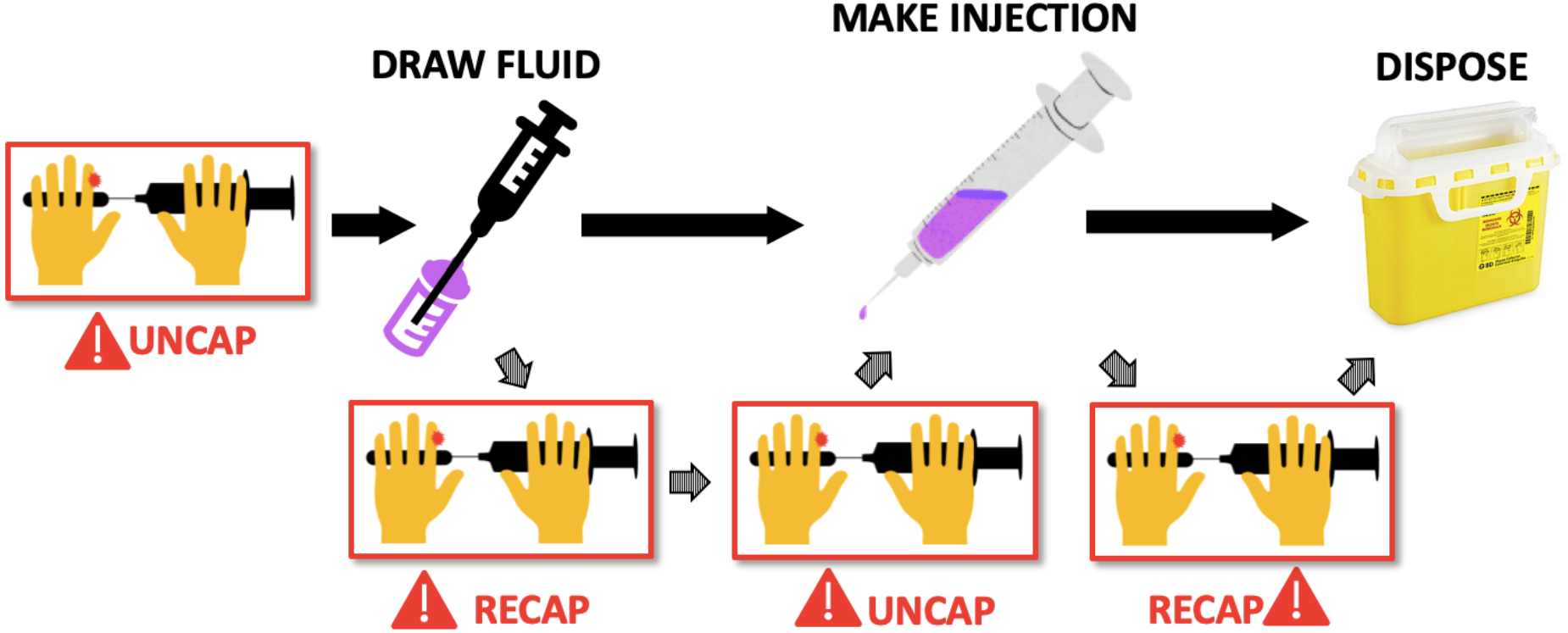
Needle-use workflow with risk event points. During the preparation, use and disposal of an injection needle, the needle-user is at risk of receiving an NSI up to 4 times from uncapping and recapping alone (13).

NSIs are a concern worldwide in both developed and underdeveloped countries; it is estimated that healthcare workers in developed countries who suffer a needlestick injury and contract hepatitis C make up about 2-4% of the total number of new hepatitis C cases each year (7). In North America, the Centre for Disease Control and Prevention reports that approximately 385,000 sharp-related injuries are recorded every year among healthcare workers (3), contributing to the worldwide total of more than 2 million NSIs experienced by healthcare workers annually (2). This roughly translates to each healthcare worker receiving on average 1.72 NSIs per year (1). According to Bouya et al. (2020), NSIs are twice as likely to occur in underdeveloped countries in the Eastern Mediterranean Region (EMRO) without implemented legislation to reinforce the use of safety-engineered medical devices (SEDs) and without adoption of the Needlestick Safety and Prevention Act in healthcare facilities, than in developed countries (2, 12).

Outside of healthcare, data on NSIs in veterinary practice, academic research, and animal research are sparse; more than 90% of academic research conducted on NSIs is derived from the healthcare sector, 10% from the veterinary sector, with minimal data on NSIs in research environments (13). This article aims to analyze a sample survey population of workers in workplaces using needles in British Columbia, investigate areas in need of more research attention, discuss trends on NSIs across multiple fields, summarise the consequences of NSIs including long-term social, financial, psychological, and health effects of NSIs in healthcare, veterinary, and research environments, and to address the recent impact of COVID-19 on NSIs.

## Methods

For the review component of this article, a non-exhaustive search of peer-reviewed scientific articles and literature reviews was completed using PubMed and OVID/Medliner (Fig. 3) with the following search terms: (“needlestick injuries” OR “needle stick injuries” OR “needle-stick injuries” OR “needlestick injury” OR “needle stick injury” OR “needle-stick injury” OR “needle injuries” OR “needle injury”) AND (“Communit*” OR “Military medicine” OR “Veterinary medicine” OR “Veterinar*” OR “Hospital” OR “Medical clinic” OR “Healthcare” OR “Psychological Impact” OR “Behavioral Impact” OR “Physical Impact” OR “Research laboratory”) with and without (“COVID19” OR “COVID-19” OR “COVID”) within the date range of 1998-2022, retrieving 1,437 results to answer the question: “What is the prevalence of NSIs in healthcare, veterinary care, research and community or military environments in developed countries?” We limited this review to research on human healthcare, veterinary care and research subjects only and included English language-based international peer-reviewed articles, online reports, electronic books, and press releases, using the following PRISMA inclusion criterion: 1. Studies that investigate needlestick injuries in healthcare facilities, community care settings, research facilities, or veterinary facilities. 2. Studies that report on the prevalence, incidence, risk factors, or consequences of needlestick injuries. 3. Studies that describe interventions or strategies to prevent or manage needlestick injuries. 4. Articles published in English. Exclusion criteria included: 1. Studies that are not related to needlestick injuries. 2. Studies that investigate other types of sharps injuries, such as cuts or puncture wounds. 3. Studies conducted in lower-income countries or the Global South. 4. Studies of needlestick injuries in other non relevant contexts (ie: farms, safe injection sites). 5. Studies not published in English. 6. Review articles. See Fig. 4 for results of search strategy. We also applied a snowballing search methodology using the references cited in the articles identified in the literature search. For the snowballing search, articles were limited to those reporting NSIs, i.e, percutaneous or sharps injuries experienced by workers employed in healthcare facilities, veterinary or research facilities, and articles analyzing the cost and consequences of NSIs in the workplace. Each identified item was assessed for relevance by a member of the study team and final calls were made by the senior author. This review is intended to summarize relevant and recent literature on NSIs in the workplace, present current population-based metrics, and provide context for the sample survey population of workers in workplaces using needles. For this study, a publicly available sample population dataset was analyzed from British Columbia (n=30) on workplace needlestick injuries. All data were anonymized and decoupled from any personal information. The Canadian Center for Occupational Hazard and Safety, World Health Organization (WHO), and National Institute of Environmental Health Sciences (NIEHS) guidelines were reviewed. Additionally, University of British Columbia (UBC) Workplace Safety Incident and WorkSafeBC databases were searched using Freedom of Information and Protection of Privacy Act (FIPPA) requests.

**Figure 3.**
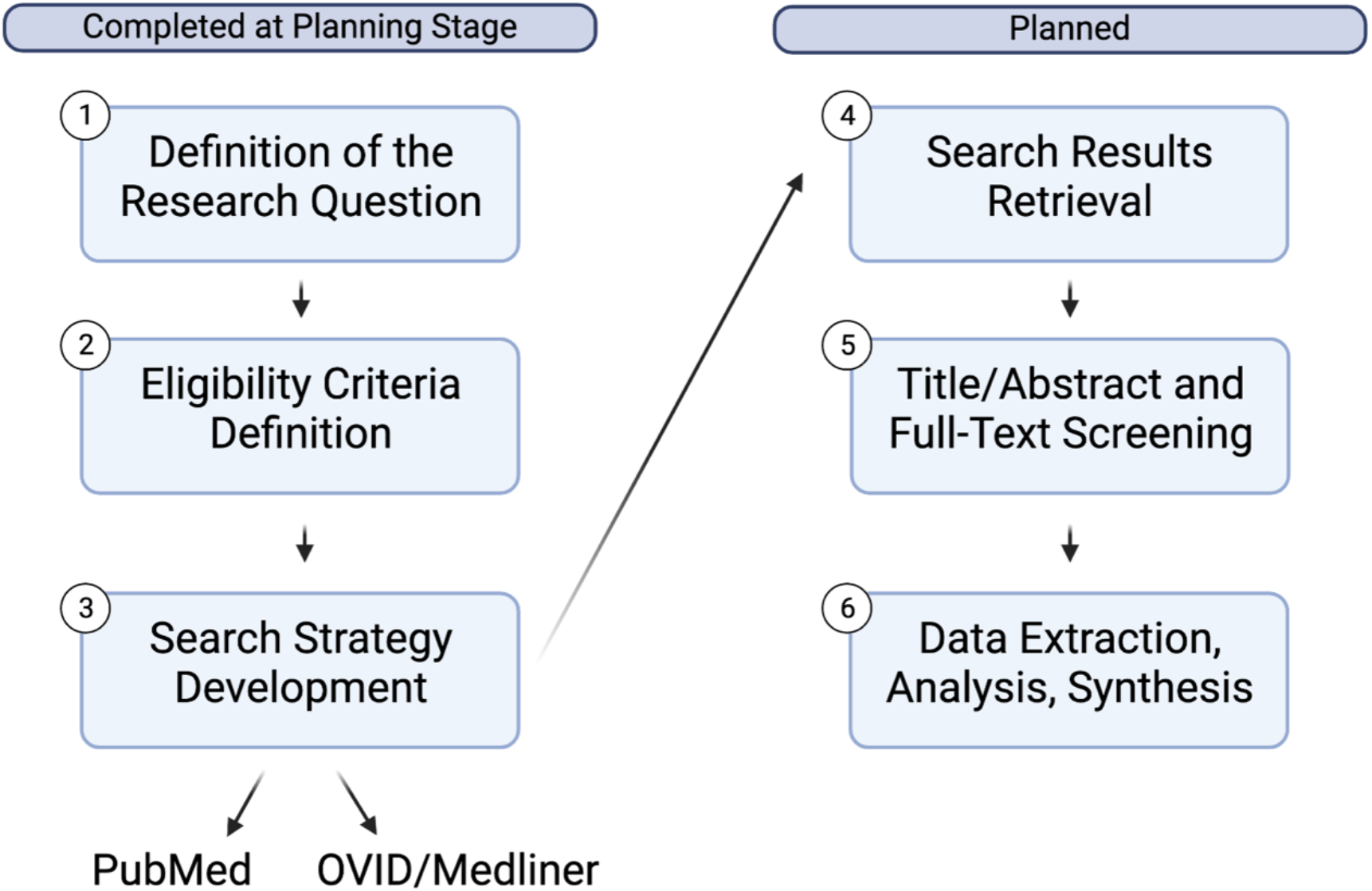
Systematic review protocol development phases used to undertake the review process.

**Figure 4.**
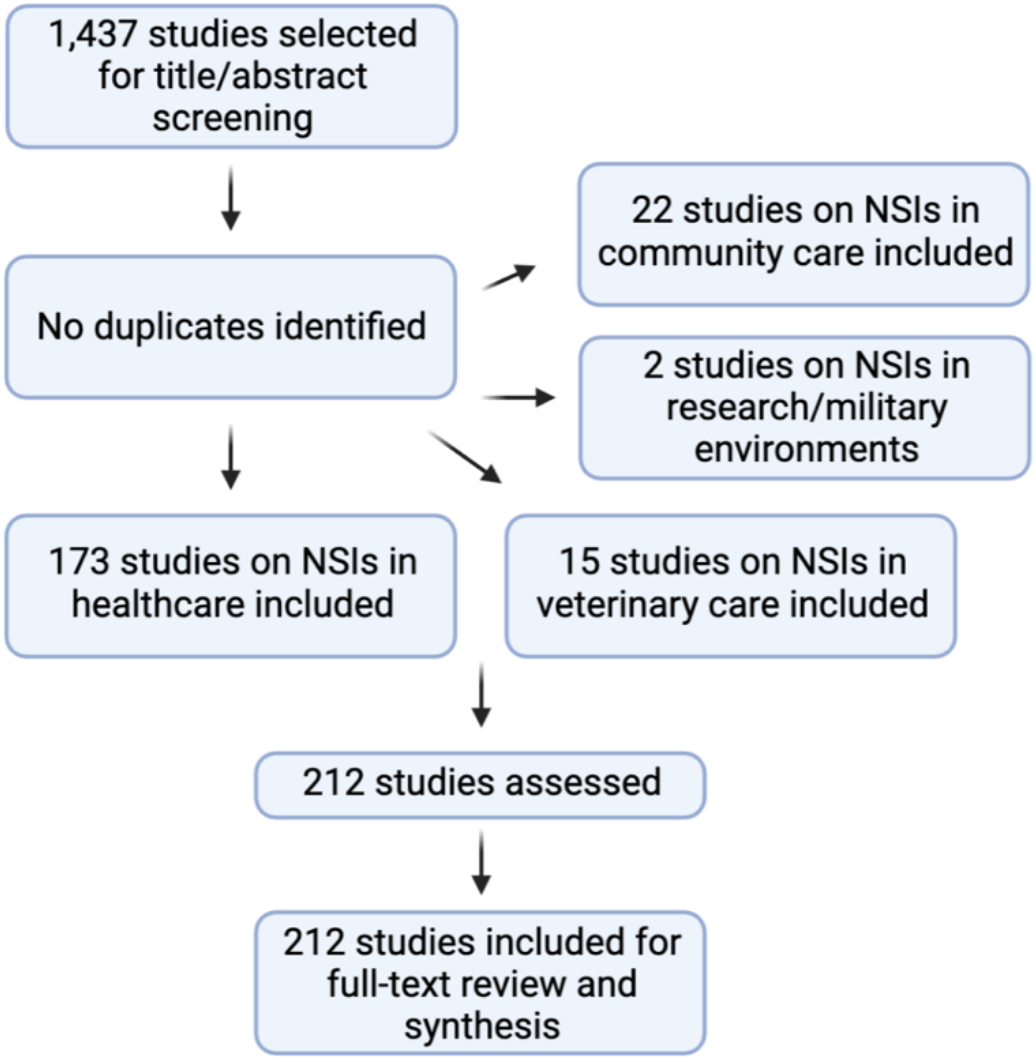
Study stratification and review according to PRISMA guidelines. Inclusion criteria: 1. Studies that investigate needlestick injuries in healthcare facilities, community care settings, research facilities, or veterinary facilities. 2. Studies that report on the prevalence, incidence, risk factors, or consequences of needlestick injuries. 3. Studies that describe interventions or strategies to prevent or manage needlestick injuries. 4. Articles published in English. Exclusion criteria: 1. Studies that are not related to needlestick injuries. 2. Studies that investigate other types of sharps injuries, such as cuts or puncture wounds. 3. Studies conducted in lower-income countries or the Global South. 4. Studies of needlestick injuries in other non relevant contexts (ie: farms, safe injection sites). 5. Studies not published in English. 6. Review articles.

## Results and Discussion

### Causes of NSIs and the Impact of COVID-19

A primary cause of NSIs in healthcare, veterinary and research environments is the unsafe recapping of needles, representing more than 25% of all NSIs in some environments (14-18). Although strongly advised against by OSHA, recapping remains common in needle usage, particularly in the veterinary and research fields (19). Previous studies have shown that in veterinary practices, up to 84.3% of veterinarians recap their needles and 74.2% of veterinarians have reported that recapping led to at least one workplace NSI (8, 20-25). Similarly, up to 90.9% of medical technologists in hospital laboratories with similar working conditions as research laboratories experienced NSIs, and reported that 24.8% of NSIs resulted from carelessness during recapping or uncapping (25). Another study of healthcare workers reported a high prevalence of NSIs among registered nurses who recap needles (26). The high prevalence of recapping needles in healthcare, veterinary and research environments, despite risk of injury and recommendations against recapping, indicates that a method to facilitate recapping safely is needed rather than efforts to completely abolish recapping.

In support of this, a study conducted by Wicker et al. (2014) surveyed medical staff and healthcare students at a hospital on their experiences with NSIs. The study examined causes and possible preventative measures for NSIs, infectious status of the patient, and whether the NSI was a concern to the worker (27). The study results revealed that the main factors leading to NSIs were worker stress or user fatigue from performing high volumes of injections, where more than 80% of respondents (with a majority who care for infectious patients) were concerned about the consequences of NSIs, and about 50% of respondents reported that NSIs could have been prevented by improving workplace safety, such as more use of adequate protective equipment and better work routines (27). Similarly, a survey conducted by Weese and Faires (2009) on veterinary technicians revealed that despite receiving adequate needle-handling training, high-risk needle-handling practices are still commonly used in veterinary clinics, including manual recapping, “one-handed scoop” methods, and uncapping with teeth (20). Taken together, these findings demonstrate that currently recommended safe alternative needle-handling practices are not effectively being adopted in many workplace settings, with expediency and familiar techniques trumping safety protocols even in the face of considerable data on the impact of NSIs.

Factors contributing to increased risk of NSIs in healthcare were investigated in a 2003 case-crossover study, which revealed that a majority of NSIs were caused by rushing (48%), followed by distraction (35%), fatigue (28%), and anger (12%) (28). Rushing, distraction and fatigue are often a result of the fast-paced nature of healthcare environments, and can be exacerbated by staffing shortages, repetitive task fatigue, and worker burnout. Therefore, strategies to alleviate work pace, improve staffing shortages, and implement SEDs to reduce risk and fatigue while using needles could help reduce NSIs. NSI research is underpowered and underrepresented in non-healthcare settings, representing less than 18% of all NSI-related studies, and research environments representing less than 1% of all studies captured in this review (Fig. 4).

During mass vaccination rollouts to combat the 2009 H1N1 Pandemic, incidences of NSIs were five times greater than levels before the 2009 H1N1 Pandemic (29). The prevalence of NSIs is likely to increase at a similar rate due to the ongoing COVID-19 pandemic as the world attempts to vaccinate 7 billion people and provide rolling booster shots to the population. Additionally, healthcare workers from diverse specialties are required to wear Personal Protective Equipment (PPE) as COVID-19 has high transmissibility while caring for an overwhelming number of patients under extremely stressful conditions. Excessive PPE may lead to poor vision and mobility, and coupled with the stress of an increased workload, performing repetitive medical procedures puts health care workers at a higher risk of NSIs and other potential health hazards (30). This problem is exacerbated by surges in the number of COVID-19 patients, which further taxes overworked and PPE-overburdened healthcare workers (31). Based on a recent study comparing 2019 (pre-pandemic era) to 2020 (pandemic era) NSI prevalence, nurses experienced more NSIs than doctors and interns (50.24% versus 16.8% and 47.4%), and the frequency of NSIs increased during the pandemic (31). Due to the current pandemic, there is more emphasis for institutions to revise and implement standard precautions of NSIs and safe needle-handling practices, to enhance performance and knowledge, and to protect personal well-being among health care workers (2).

### Psychological and physical effects of NSIs

According to WHO statistics, NSIs cause 1,000, 16,000, and 66,000 cases of HIV, HCV, and HBV respectively, among healthcare workers every year (32). Healthcare workers who experienced NSIs also exhibited psychological impacts, including higher levels of anxiety and depression. This is supported by a study conducted in Korea where stress, anxiety, and depression scores of workers were measured using information from a routine examination prior to the study and utilizing the Beck Depression Inventory (BDI), Hamilton Anxiety Scale (HAM-A), and Perceived Stress Scale (PSS) (33). To summarize, the BDI is used for self-reported inventory that measures the severity of a depressive mood where a higher score indicates higher severity, the HAM-A is a 0-14 scale used to quantify anxiety symptoms, and the PSS is a self-reported questionnaire where a high score indicates greater stress, but does not take into account factors such as sex, age, and education (33). The results showed that 71.1% of healthcare workers experienced NSIs (mostly comprising female nurses), and NSI-affected healthcare workers exhibited higher PSS and BDI scores after an injury as compared to colleagues who had not received an NSI (33). Similarly, a study conducted on nurses caring for patients with diabetes who self-reported their past year’s experiences on NSIs, revealed that 60% of 110 nurses who received at least one NSI from blood draws or malfunction of needle safety disposal equipment reported enhanced fear of needles two weeks after exposure, with about 41.8% also feeling anxious, depressed, or stressed post-injury (34). A similar study was conducted by Gershon et al. (2000) on NSI-affected healthcare workers reported increased feelings of anxiety (53%), insomnia (18%), depression (13%), loss of appetite (10%), sleepiness (10%) and frequent emotional breakdowns (10%) after NSIs (35).

There also appears to be a lack of support programs for workers receiving NSIs at the time of exposure, and minimal anticipatory discussions on what one should do when exposure occurs. This can exacerbate feelings of anxiety and depression and result in further deterioration of the recipient’s mental health, leading to extended work absences and ultimately departure from the profession. A study conducted by Greens & Griffiths (2013) tested the duration and severity of NSI-induced psychiatric disorders compared to non-NSI-induced psychiatric disorders (for instance, trauma from road traffic accidents) through individual interviews with a psychiatrist (36). Participants were assigned a Beck Depression Inventory (BDI) score, and BDI scores in NSI patients indicated moderately severe depression (independent of age and sex) which decreased over time, but psychiatric illness lasted 1.78 months longer per every month an NSI patient waited for serology results (36).

While waiting for the serology results after exposure, 76% of NSI patients with adjustment disorders (AD) did not feel reassured by occupational health or emergency staff and believed they were infected with a virus, despite all NSI patients eventually receiving negative serology results (36). The lack of post-exposure care and instructions after an NSI is emphasized by Reutter and Northcott (1995), where nurses who were exposed to HIV via NSIs while caring for AIDS patients displayed their own unhealthy coping mechanisms: minimizing the effect of exposure by disinfecting and “bleeding” out the affected area, reducing a sense of vulnerability by avoiding situations that aroused fear and HIV testing, selective disclosure to others to avoid social rejection, and assigning meaning on how NSIs would impact their lives (37). A recent case report by Worthington et al. (2006) analyzed the behaviors of two separate healthy healthcare workers who received NSIs from the same HIV patient a year apart (38). Despite receiving negative test results for HIV and receiving treatment with antiviral drugs following exposure, both healthcare workers suffered from PTSD with symptoms including depression, anxiety, insomnia, nightmares, and attempts to return back to work that triggered panic attacks (38). Worthington et al. (2006) also highlighted in their case report that workers exposed to HIV infection via NSIs were more likely to be diagnosed with PTSD, and were not able to return to work as they required ongoing psychiatric care (38).

The consequences of receiving NSIs in research and veterinary environments differ from the healthcare workplace and may be more complex and detrimental to the user’s health. Depending on the working environment, researchers regularly handle high concentrations of toxic chemicals, drugs, cancer stem cells, and bloodborne pathogens such as hepatitis B (HBV), hepatitis C virus (HCV), human immunodeficiency viruses (HIV), and other extremely infectious or deadly viruses (23, 32, 39-43). There have also been case reports of dengue, malaria, leishmaniasis, and meningitis being transmitted by laboratory infection due to NSIs (41-43). Consequences of exposure to these agents via NSI range from illness, severe tissue damage, increased cancer risk, auto-immune events, limb amputations, and even death (39-40). For instance, an animal care specialist, who accidentally self-injected himself with a small amount of bovine vaccine, experienced a loss of sensation in his finger and recurrence of lymphangitis (44). Despite early surgical treatment, amputation was required (44). In another case, a student researcher injected himself with dichloromethane, resulting in significant tissue injury (40). Additionally, 2 out of the 4 reported cases of laboratory-associated Zika virus in the United States from 2016 to 2019, were linked to NSIs (45).

NSIs can also occur in non-healthcare settings. In 2009, a California study of injection drug users (IDUs) found that IDUs accounted for approximately 9% of 48,100 new HIV infections, 15% of 43,000 new HBV infections, and 44% of 17,000 new HCV infections in the U.S based on surveillance data (46). Thus, the risk of NSIs and the risk of blood-borne pathogens (BBP) transmission is also more likely to occur in areas populated with IDUs where used and discarded needles are found in public areas. NSIs can also occur in community settings; a study by Kordy et al. (2017) assessed the effects of discarded needles in children aged less than 19 years old, in community areas (47). Out of the 66 community-based exposures, 71.2% were from needles contaminated with BBPs where exposures occurred in outdoor communities such as parks (45.5%), schools (30.3%), homes (15.2%), and out-patient clinics (9%) (47). The results also showed that only 15 out of 22 children completed the full course of HIV post-exposure prophylaxis, and only 41.2% of previously unvaccinated children completed the HBV vaccine series post-exposure (47). Additionally, the study found that only 60.6% of NSI patients have gone through and received adequate follow-up testing (47). Similarly, a review analysis of community-acquired NSI (CANSIs) records from 1991 to 1996 for children under the age of 20 in the Canadian Hospitals Injury Reporting and Prevention’s surveillance network showed that about 78% of CANSIs were classified as high-risk and presumably and likely a result of discarded needles from IDUs, 6% of CANSIs as intermediate-risk, and 12% of CANSIs as low-risk (48). About 77% of the injuries involved the needle being picked up between ages 0 to 9 years old and these NSIs occurred in outdoor settings, mostly in parks (21%) (48). Vaccines for HBV were given to 76-78% of the children as only 1.7% of children infected were immunized against this virus, since immunization is generally given to children in grades 4 or higher during the time of the study (48). Hence, revising the administration of HBV to early infancy might help eliminate the need to administer urgent HBV vaccines and follow-up testing as based on previous studies, not all patients were able to complete their post-NSI exposure treatments. Additionally, programs aimed to reduce environmental contamination of contaminated BBP needles by the creation of public sharp disposal bins, and enhanced public health educational interventions about the potential risks of used needles that target parents, students, and school staff may help reduce the risk of NSIs and lower transmission of HBC, HCV, and HIV.

NSI incidence for healthcare workers in the in-home healthcare setting are rapidly growing due to the popularity and convenience of in-home healthcare services. However, data in this aspect is limited and recent data is needed to determine the severity of NSIs in-home care environments. Haiduven et al. (2004) studied and analyzed NSIs and blood exposure reports from 1993 to 1996 obtained from three home health care agencies in San Francisco Bay (49). Out of 52 exposures, 48 were from needlesticks and 92% of these injuries were sustained by home care nurses (49). Injuries occurred prior, during, and after needle disposal (23%), manipulating intravenous lines (17.3%), improper disposal (15%), and during or after blood draws (13.5%) (49). Another study by Perry et al. (2001) examined data from the Exposure Prevention Information Network in 1993 to 1998 from home health settings to determine the pattern and frequency of NSIs (50). Almost half of NSIs (48%) occurred during the disposal of and withdrawal of blood using hollow-bore needles, disposable syringes, phlebotomy needles, lancets, and winged steel needles (50). Interestingly, Perry et al. (2001) stated that 87% of home care nurses reported sustaining an NSI compared to 65% of nurses who reported sustaining an NSI in hospitals (50). Hence, the data shows that home health care providers have a higher frequency of NSI compared to their hospital counterparts. The high frequency of NSI is likely due to the unique environment home care nurses work in, and hence, needle safety devices specifically tailored to home settings are needed (50). There has been minimal research on interventions that could help mitigate NSIs in veterinary care, research, and in-home care, but possible solutions are to expand NSI education and prevention programs outside of medical facilities, select safer sharps devices and improve safety training in inexperienced personnel and needle-users in high-risk environments.

### NSIs Reporting

Although most workplaces have protocols for reporting NSIs, reporting habits remain very poor, particularly among veterinary and research employees. A recent review showed that despite a high incidence of NSIs in a sample of veterinary clinics, reporting was extremely low (51). A field study from 2013 to 2015 at Toronto East General Hospital (TGEH) involving 350 medical students, residents, and postgraduate fellows revealed that about 25% of respondents had experienced at least one NSI recently, and only 28 out of 88 (32%) recent NSIs were reported (52). Some reasons for underreporting included the perception that NSIs posed an insignificant risk to respondents’ health, lack of resources and support after experiencing an NSI, and injury stigmatization (52). Similarly, studies show that up to 94% of NSIs are not reported by healthcare workers, and up to 99% of NSIs in veterinary care are not reported, with common reasons for non-reporting being belief that the needle was sterile, lack of concern for NSIs, the perception that reporting workplace injuries are inconvenient or time-consuming and desire to avoid associated stigma (51,53-60). Oftentimes, poor behaviour modelling exacerbates this issue, as supervisors do not report their own NSIs and may advise or imply that their employees should not report NSIs as well to avoid completing additional paperwork. This has been linked to heightened stress, trauma, and feelings of ostracization among workers experiencing NSIs (34, 38).

Globally, healthcare workers receive an average of 1.72 sharps injuries per healthcare worker per year (1). We performed a study on a sample population of workers using needles across healthcare (5%), emergency healthcare (22%), veterinary care (46%), and animal research (27%) (Fig. 5A). Our study indicates that a vast majority of workers have experienced a needlestick injury, upwards of 90% of workers in certain workplaces, including animal research and veterinary care (Fig. 5B), that a significant proportion do not report (Fig. 5C), and that over half of these needlestick injuries occur due to uncapping or recapping needles (Fig. 5D-F).

**Figure 5.**
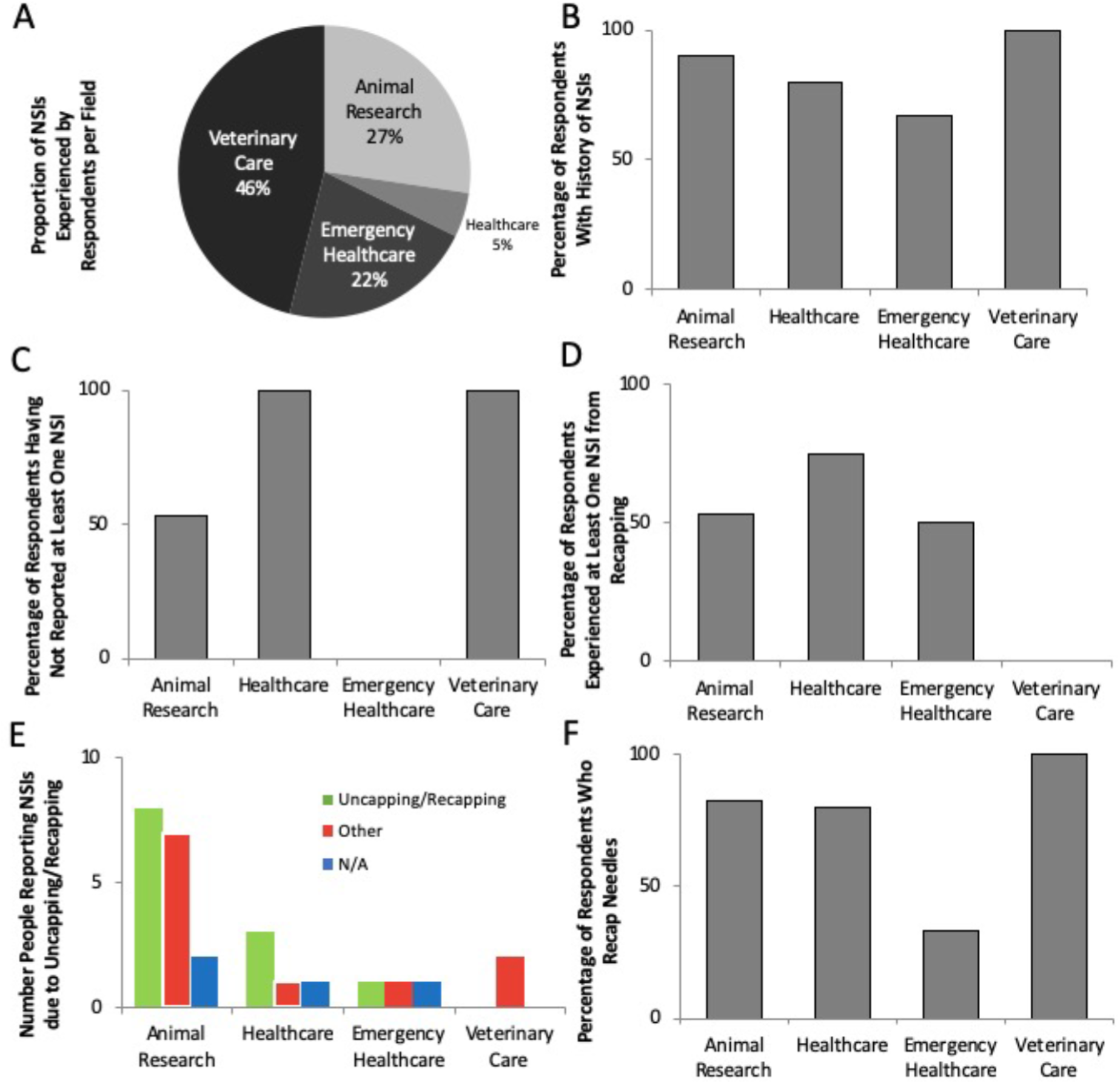
Needle-stick injury data from workplaces across BC. (A) Total proportion of respondents by field. Percentage of respondents (A) reporting a history of NSIs in the workplace, (B) reporting recapping as regular practice, (C) admitting to not reporting at least one NSI, (D) reporting at least one NSI from recapping from a sample of workers in BC. Number of respondents reporting/ not reporting NSIs. (F) Proportion of respondents who recap needles. (n = 30).

To verify this data, we performed a sample data extraction of workers in two representative populations: the entirety of the Canadian province of British Columbia (BC), and the University of British Columbia (UBC). FIPPA data obtained from WorkSafeBC indicates that NSIs in BC make up approximately 1% of all workplace compensation claims in recent years, although this data is likely skewed by underreporting, based on excessive rates of underreporting demonstrated in academic literature, and due to WorkSafeBC’s database including many workplaces that do not use needles in assessing the proportion of injuries caused by needles in the workplace. Despite this, in a ten-year reporting period, WorkSafeBC still found multiple instances of infectious disease transmission and injury, including instances of HIV infection as a result of NSIs. As expected, NSIs were found to be more prevalent in workplaces with high needle-usage frequencies: data from the UBC Workplace Safety Incident database indicated that NSIs comprised more than 10% of all workplace injury reports at UBC, an institution with several research facilities and hospitals (61).

### Financial effect of NSIs

The consequences of NSIs may lead to work loss and significant financial costs to the employer and to the workers themselves (62-63). In the US, each NSI on an individual basis costs on average $596 USD; these results are very similar to FIPPA data obtained from WorkSafeBC on NSIs, with an annual national cost in the USA of $188.5 million USD (direct medical costs including healthcare visits, drugs and medical treatment costs of $107.3 million USD and indirect costs from lost-work, loss of productivity and worker retention-loss of $81.3 million USD) (64). Similarly, in Europe, NSIs have an estimated economic burden of $300 million EU per year in England and Wales, and Germany estimates an annual cost of $30 million EU per year in direct costs from reported NSIs and estimates more than $133 million EU annually in direct costs from unreported NSIs (65). Additionally, indirect costs from lost productivity associated with time taken to make reports, receive treatments, recover from emotional distress and anxiety, and long-term health, litigation, or employment effects are typically left out of calculations of NSI costs due to a focus on more easily tabulated immediate post-exposure medical and work loss costs. This likely increases the cost of NSIs significantly beyond these conservative projections. The financial burden to the economy caused by NSIs is expected to increase over time until more cost-effective, user-friendly and efficacious solutions including more efficacious SEDs are established to significantly lower the incidence of NSIs in the workplace (66).

### Prevention and control of NSIs

Current NSI prevention centers primarily around SED implementation and is divided into three categories: behavioural directives (including safe needle use and disposal practices), engineering controls, and risk-reduction tools (including safety needles and safe needle disposal units). Behavioural directives, consisting of training and guidelines against recapping, are not always strictly complied with by workers due to conflicts between safety protocols (theory) and procedure protocols (practice). As well, the need to recap to maintain sterility or avoid an accidental poke while transferring the needle and syringe to another room to perform an injection often necessitates unsafe recapping behaviours. Engineering controls, such as safety needles, are cumbersome, difficult to use, and generally avoided by workers in favour of regular needles (67). Most safety needle models are only designed to recap once, making this feature unfeasible to use until the needle is ready to be disposed of, and requiring workers to rely on unsafe recapping practices while using the needles. Safety needles are also environmentally unfriendly and ten-fold more expensive than regular needles, making them an unworkable, underused, or unaffordable solution in many work environments (68). Additionally, despite their name, safety needles have still been reported to cause NSIs (67-68). Currently, single-handed risk-reduction tools are rare, and include needle-uncappers like NeedleSafe II. These safety engineered devices have low uptake possibly due to compatibility limitations, limited workplace adaptability, and ergonomic issues. Other interventions that have been studied for their effectiveness and impact on reducing NSIs in healthcare settings including double gloving, utilization of blunt needles, and educational training, suggesting that increased safety measures and educational training programs can reduce NSIs in the workplace, particularly among vulnerable groups such as medical and nursing students, as well as workers performing repetitive injections or injections under duress during emergency surgery (69-70). However, based on the current prevalence of NSIs and the options offered in the current market for NSI preventatives, an affordable, versatile, and efficient NSI device is still needed.

## Conclusion

In conclusion, NSIs affecting workers in healthcare, research, and veterinary environments have complex and long-term health, psychological and financial impacts. As concerns over NSIs continue to grow, and in order to protect the overall safety of needle users while minimizing economic losses, a new generation of affordable, versatile, and efficacious NSI-prevention devices are needed, with corresponding SED and behavioural training regimens to reduce NSIs in the workplace. With the increase in vaccinations due to the COVID-19 pandemic, prevalence of NSIs, particularly in developed countries, is expected to rise, contributing to worker stress and burnout. This study and literature review may be used as a basis to plan for additional solutions to reduce NSI rates in the workplace, including the implementation of SEDs, establishing clear and uniform policies or awareness programs for NSI management in hospitals, clinics and research facilities, and performing periodic testing on staff knowledge of standard protocols for NSI prevention.

## Data Availability

Source data were openly available before the initiation of the study : https://www.decaprandd.com/decap-whitepaper. All data produced in the present study are available upon reasonable request to the authors.

## Authors’ contribution

Literature review: SYSL, JM

Tables and figures: SYSL, IN

Writing: SYSL, IN, JM

Review and comments: SYSL, JM

## Funding and Ethics

No specific funding was allocated for this research. Source data were openly available before the initiation of the study : https://www.decaprandd.com/decap-whitepaper. All data produced in the present study are available upon reasonable request to the authors.

